# Clinical Profile, Aetiology, and Temporal Trends of Hospitalized Heart Failure Patients in a Private Tertiary Hospital in Sierra Leone (2021 – 2025)

**DOI:** 10.64898/2026.06.06.26355075

**Authors:** James Baligeh Walter Russell, Mohamed Smith, Yakubu Alhassan, Joshua M Coker, Edries Tejan, Kumar Bharat, Meena Kumari, Othman Z. Mahdi, Durodami Radcliff. Lisk

## Abstract

**Background:** Heart Failure (HF) is a complex clinical syndrome of growing public health concern in sub-Saharan Africa, yet data from Sierra Leone are absent. The aim of the study is to characterize the clinical profile, etiological and temporal trends of hospitalized HF patients at Choithrams Memorial Hospital (CMH), in Freetown, Sierra Leone, to inform specific management strategies.

**Methods:** This single-centre, retrospective observational cohort study analysed data on HF patients (≥ 18years) admitted at the CMH between January 2021 to 31 December 2025. The clinical definition of HF was based on the Framingham criteria and the European Society of Cardiology (ESC) guidelines, including standard echocardiographic parameters. All variables, including patients’ demographics, HF phenotype, aetiology, medical history, and hospital outcomes, were extracted from the digital record. Non-parametric tests, multivariable logistic regression to identify variables associated with aetiology, Wilcoxon rank-sum test to compare groups, and Kruskal-Wallis test to analyse trends over time were utilised.

**Result:** A total of 765 patients were included in the study, with a median age of 53 years (IQR 42-61) and male predominance of 55.3%. Patients with recurrent HF (60.9%) were more common than those with *de* novo HF (39.1%), were older (54 years vs 53 years), had a higher comorbidity burden (34% vs 4%, p < 0.001), and presented with a cold-wet hemodynamic profile (18.4% vs 8.4%, p < 0.001). HFrEF (61.3%) was the most predominant phenotype, though HFpEF increased with age. Dilated Cardiomyopathy (37.0%), Hypertensive Heart Disease (31.2%) and Valvular Heart Failure (17.1%) were the leading etiologies, while ischemic heart disease (6.3%) was relatively uncommon. A majority of the patients were referred (77.9%), and 50.8% presented with NYHA IV. The strongest independent predictor for HF was hypertensive heart disease [AOR = 17.81; C.I 95%: (3.13-48.76), p <0.001]. An analysis of the trends in etiologies and demographics over the five-year period demonstrated no significant changes (all p-values > 0.05 for age, sex, aetiology, and most comorbidities).

**Conclusion:** HF affects the younger adult population in Sierra Leone and is mainly caused by DCM and HHD. The late case presentations, the high prevalence of recurrent HF, and the associated high burden of comorbidities emphasize an urgent need to develop and implement improved strategies for the prevention, early detection, and long-term management of HF within Sierra Leone’s healthcare system.

## 1.0 Introduction

Heart failure [HF] is a complex clinical syndrome of major public health concern, affecting an estimated 64 million people worldwide. [1-3]. Although HF has been studied extensively in high-income countries, its epidemiological characteristics differ in developing countries. These differences are mainly due to deficiencies in the management of chronic cardiovascular disorders within health systems. [4-7]. Studies from major African heart failure registries, including THESUS-HF [7] and INTER-CHF [2], have also provided valuable insight into these regional inequalities.

In sub-Saharan Africa (SSA), the prevalence of heart failure is increasing, a trend attributable to the ongoing epidemiological transition from communicable to non-communicable diseases. This transition is accompanied by rising incidence rates of hypertension, diabetes mellitus, and obesity [3,4,6,8]. Peculiar to the African setting, a younger age demography has been reported, which is attributed to non-ischemic aetiology, particularly untreated hypertension, rheumatic heart disease and cardiomyopathy [3,9]. Additionally, the substantial burden of infectious diseases may contribute to the region’s distinct clinical profile of heart failure [7,9]. By contrast, in high-income countries, heart failure most frequently results from ischemic heart disease, primarily in older adults [4,8]. These significant differences underline the importance of developing strategies and interventions that are designed to the unique causes and realities of heart failure in sub-Saharan Africa [5,10]. In the West African subregion, Heart Failure is a common, life-threatening condition among other cardiovascular diseases, representing about 6 to 10% of all hospital admissions [9,11,12].

Sierra Leone is a low-income country in West Africa, facing considerable challenges in its health system, which is heightened by the aftermath of a decade of civil war and the Ebola outbreak. Data on the clinical characteristics and aetiology of heart failure in Sierra Leone are absent, and most information on this condition is extrapolated from sub-regional studies. Therefore, these data may not fully reflect the national picture of the disease, given disparities in genetic, environmental, and healthcare availability.

A comprehensive understanding of the specific characteristics of the HF population in Sierra Leone constitutes an important initial step towards developing effective, context-appropriate public health interventions and clinical management protocols. Hence, there is an urgent need to characterise the risk factors and aetiology of heart failure in Sierra Leone. The aim of the study is to describe the clinical profile, aetiology, and trends of hospitalized heart failure patients at the Choithrams Memorial Hospital, Freetown, Sierra Leone, over a period of five-years.

## 2.0. Patients and Methods

### 2.1. Study area, design and Ethics approval

This was a single-centre, retrospective observational cohort study conducted at Choithrams Memorial Hospital (CMH), an affiliated tertiary healthcare facility of the University of Sierra Leone Teaching Hospital Complex in Freetown, Sierra Leone. A 5-year period (January 2021 to December 2025) was selected to assess the study’s reliability regarding sample size, generalizability, and temporal trends in heart failure patterns. CMH is a 60-bedded hospital, providing tertiary healthcare services to the urban community, with particular focus on advanced diagnostic evaluations and specialized medical treatment.

The hospital has a total of 60 beds, providing tertiary-level healthcare services to the urban community, with a particular focus on advanced diagnostic evaluations and specialized medical treatment. CMH provides a 24-hour emergency service in addition to outpatient specialist clinics, handling an average of around 40 patients each month. While most referrals were from the Western Area, the hospital also receives patients from district hospitals and other private health facilities throughout the country, indicating its extensive referral network. The study was approved by the Sierra Leone Ethics and Review Committee.

### 2.3. Eligibility criteria, procedure, and data collection

The study enrolled hospitalized patients if they were ≥ 18 years, met the 2021 ESC diagnostic criteria for HF, Framingham criteria and had a complete echocardiogram (M-mode, 2-dimensional and Doppler measurement) performed during the index admission [13,14]. A Cardiologist and a certified Echocardiographer performed all echocardiographic studies by using a standard GE 2D Echocardiogram with a 2.5 MHz probe. Patients were categorized into three heart failure phenotypes based on Left Ventricular Ejection Fraction: Heart Failure with reduced ejection fraction (HFrEF): LVEF ≤ 40%; Heart Failure with mildly reduced ejection fraction (HFmrEF): 41% ≤ LVEF ≤ 49%; and Heart Failure with preserved ejection fraction (HFpEF): LVEF ≥ 50%.

The aetiologies of HF were assigned by the Cardiologist based on clinical, echocardiographic, and electrographic findings. Patients with inadequate records of less than 80% of key variables and with primary conditions unrelated to heart failure were excluded.

As a retrospective study utilising previously collected clinical data, the sample size calculation was not performed, and hence informed consent was not obtained. This approach is in keeping with ethical principles and guidelines that typically waive the consent requirement for pre-existing data. A data extraction tool was used to collect medical information for the study. All case records were anonymised by assigning serial codes, and the extracted data were processed with strict confidentiality.

The data were extracted by using a standardised collecting form covering the following: 1. Demographic Characteristics: sex, age, occupation, body mass index (BMI), and number of comorbidities. 2. Medical History: Family history and specific cardiovascular family history. 3. Medical findings such as blood pressure, mean arterial pressure, heart rate, medical history of hypertension, coronary artery disease, left ventricular ejection fraction (LVEF), left ventricular end-diastolic diameter (LVDD), echocardiographic findings, laboratory test results, length of hospitalization, in-hospital outcomes, and classifications of heart failure (reduced, mildly reduced, and preserved). Clinical presentation (self-presentation or referral), aetiology of heart failure, cardiovascular risk factors, NYHA (II-IV)classification, and hemodynamic profile (warm-dry, warm-wet, cold-wet, cold-dry) were also documented.

#### 2.3.1 Study participants and procedure

Patients were categorized into three heart failure phenotypes based on Left Ventricular Ejection Fraction: Heart Failure with reduced ejection fraction (HFrEF): LVEF ≤ 40%; Heart Failure with mildly reduced ejection fraction (HFmrEF): 41% ≤ LVEF ≤ 49%; and Heart Failure with preserved ejection fraction (HFpEF): LVEF ≥ 50%.

### 2.3. Statistical analysis

Stata version 19.5 (StataCorp, College Station, TX) was used to analyse the data. Descriptive statistics were used to summarize demographic, clinical, aetiological, and comorbidity profiles. Continuous variables were assessed for normality using the Shapiro–Wilk test and visual inspection of histograms and Q–Q plots.

As all continuous variables (age, systolic and diastolic blood pressure, mean arterial pressure, heart rate, and BMI) departed from normality, they were summarized as medians with interquartile ranges (IQR). Categorical variables were summarized as frequencies and column percentages. Bivariate comparisons of continuous variables between two independent groups (*de* novo vs recurrent HF; female vs male) were performed using the Wilcoxon rank-sum (Mann–Whitney U) test. Median comparisons across three or more independent groups (e.g., age categories, year of admission) were analysed using the Kruskal-Wallis test. Pearson’s chi-square were used to assess associations between categorical variables. Temporal patterns throughout the five-year study period (2021–2025) were assessed using the Kruskal–Wallis test for continuous variables and the chi-square test for categorical variables.

To identify factors independently associated with each of the five major aetiologies, namely, dilated cardiomyopathy (DCM), hypertensive heart disease (HHD), valvular heart disease (VHD), ischemic heart disease (IHD), and other aetiologies, separate multivariable binary logistic regression models were used, with each aetiology serving as a binary outcome (presence versus absence). Candidate predictors included age group, sex, BMI category, presentation circumstances, patient type (de novo versus recurrent), year of admission, cardiovascular risk factors (alcohol intake, family history of heart disease), and comorbidities (hypertension, diabetes mellitus, stroke, COPD, chronic kidney disease, anemia, cancer, thyroid disease, and atrial fibrillation).

Smoking history was excluded from the IHD model due to zero observations among smokers and from other models in which complete separation was identified. In cases where small cell counts led to unstable estimates, sparse categories were combined; for example, the 20–39 and 40–59 age groups were merged as the reference for IHD, and the overweight and obese categories were combined for IHD.

All Results from the logistic regression models are presented as adjusted odds ratios (aOR) with 95% confidence intervals (CI). Model fit was assessed using the Hosmer–Lemeshow goodness-of-fit test, and multicollinearity among predictors was evaluated using variance inflation factors (VIFs), with VIFs> 5 indicating problematic multicollinearity. Cases with missing covariate data were handled using complete-case analysis in the multivariable models. A two-sided p-value < 0.05 was considered statistically significant. Given the exploratory nature of multiple comparisons across aetiology models, no formal correction for multiplicity was applied; results were interpreted with appropriate caution.

## 3.0 RESULTS

### 3.1 Demographic and Clinical Profile

Out of the total 765 patients hospitalized with heart failure, recurrent heart failure was more common than *de* novo presentations (60.5% vs. 39.5%). The median age of the cohort was 53 years (IQR: 42-61) with a male predominance (55.3%). Patients with recurrent heart failure were older (median 54 versus 53 years; p = 0.025), with a higher proportion aged 60 years or older (32.6% in recurrent versus 25.5% in *de* novo cases; p = 0.003). No statistically significant association was observed between sex and type of patient (p = 0.650). (Table 1).

**Table 1:** Demographics, Aetiology and clinical profile of heart failure patients by type of patients.

| Characteristics | Total<br>N=765 | Do novo<br>N=302 | Recurrent<br>N=463 | P-value |
| --- | --- | --- | --- | --- |
| **Age in years, Median (IQR)** | 53 (42, 61) | 53 (38, 60) | 54 (43, 61) | 0.025<sup>w</sup> |
| **Age group, n (%)** |  |  |  | 0.003 |
| 20-39 | 162 (21.2) | 82 (27.2) | 80 (17.3) |  |
| 40-59 | 375 (49.0) | 143 (47.4) | 232 (50.1) |  |
| 60+ | 228 (29.8) | 77 (25.5) | 151 (32.6) |  |
| **Sex, n (%)** |  |  |  | 0.650 |
| Females | 342 (44.7) | 132 (43.7) | 210 (45.4) |  |
| Males | 423 (55.3) | 170 (56.3) | 253 (54.6) |  |
| **SBP (mmHg), Median (IQR)** | 119 (97, 144) | 124 (106, 149) | 114 (92, 138) | <0.001<sup>w</sup> |
| **DBP (mmHg), Median (IQR)** | 77 (66, 92) | 79 (68, 93) | 76 (65, 88) | 0.013<sup>w</sup> |
| **MAP, Median (IQR)** | 90 (80, 112) | 92 (83, 115) | 88 (77, 104) | <0.001<sup>w</sup> |
| **Heart Rate, Median (IQR)** | 84 (74, 95) | 83 (72, 92) | 85 (76, 97) | 0.005<sup>w</sup> |
| **BMI, Median (IQR)** | 22 (19, 25) | 22 (20, 25) | 22 (19, 26) | 0.930<sup>w</sup> |
| **BMI category, n (%)** |  |  |  | 0.074 |
| Underweight (<18.5kg/m<sup>2</sup>) | 152 (19.9) | 50 (16.6) | 102 (22.0) |  |
| Normal (18.5-24.9kg/m<sup>2</sup>) | 397 (51.9) | 174 (57.6) | 223 (48.2) |  |
| Overweight (25.0-29.9kg/m<sup>2</sup>) | 118 (15.4) | 43 (14.2) | 75 (16.2) |  |
| Obese (>30.0kg/m<sup>2</sup>) | 98 (12.8) | 35 (11.6) | 63 (13.6) |  |
| **Circumstance of presentation, n (%)** |  |  |  | <0.001 |
| Self-presentation | 169 (22.1) | 121 (40.1) | 48 (10.4) |  |
| Referral | 596 (77.9) | 181 (59.9) | 415 (89.6) |  |
| **Aetiology** |  |  |  |  |
| Dilated Cardiomyopathy (DCM), n (%) | 283 (37.0) | 129 (42.7) | 154 (33.3) | 0.008 |
| Hypertensive heart disease (HHD), n (%) | 239 (31.2) | 116 (38.4) | 123 (26.6) | <0.001 |
| Valvular Heart Disease (VHD), n (%) | 131 (17.1) | 72 (23.8) | 59 (12.7) | <0.001 |
| Ischemic Heart Disease (IHD), n (%) | 48 (6.3) | 27 (8.9) | 21 (4.5) | 0.014 |
| Other Aetiology, n (%) | 54 (7.1) | 10 (3.3) | 44 (9.5) | 0.001 |
| **Cardiovascular risk factors** |  |  |  |  |
| Smoking history, n (%) | 154 (20.1) | 6 (2.0) | 148 (32.0) | <0.001 |
| Alcohol intake history, n (%) | 82 (10.7) | 43 (14.2) | 39 (8.4) | 0.011 |
| Family history of Heart disease, n (%) | 178 (23.3) | 34 (11.3) | 144 (31.1) | <0.001 |
| **Comorbidities** |  |  |  |  |
| Hypertensive, n (%) | 297 (38.8) | 111 (36.8) | 186 (40.2) | 0.340 |
| Diabetes Mellitus (DM), n (%) | 162 (21.2) | 12 (4.0) | 150 (32.4) | <0.001 |
| Stroke comorbidity, n (%) | 73 (9.5) | 27 (8.9) | 46 (9.9) | 0.650 |
| COPD, n (%) | 141 (18.4) | 5 (1.7) | 136 (29.4) | <0.001 |
| Chronic Kidney Disease (CKD), n (%) | 197 (25.8) | 81 (26.8) | 116 (25.1) | 0.580 |
| Anemia, n (%) | 201 (26.3) | 92 (30.5) | 109 (23.5) | 0.033 |
| Cancer, n (%) | 19 (2.5) | 8 (2.6) | 11 (2.4) | 0.810 |
| Thyroid disease, n (%) | 51 (6.7) | 14 (4.6) | 37 (8.0) | 0.069 |
| Atrial Fibrillation, n (%) | 130 (17.0) | 34 (11.3) | 96 (20.7) | <0.001 |
| **Number of comorbidities, n (%)** |  |  |  | 0.590 |
| None | 77 (10.1) | 34 (11.3) | 43 (9.3) |  |
| 1 | 274 (35.8) | 110 (36.4) | 164 (35.4) |  |
| 2 | 414 (54.1) | 158 (52.3) | 256 (55.3) |  |
| **NYHA Functional Classification, n (%)** |  |  |  | 0.130 |
| NYHA-II | 130 (17.0) | 41 (13.6) | 89 (19.2) |  |
| NYHA-III | 246 (32.2) | 101 (33.4) | 145 (31.3) |  |
| NYHA-IV | 389 (50.8) | 160 (53.0) | 229 (49.5) |  |
| **Hemodynamic profile, n (%)** |  |  |  | <0.001 |
| Warm-dry | 89 (11.6) | 36 (11.9) | 53 (11.4) |  |
| Warm-wet | 498 (65.1) | 205 (67.9) | 293 (63.3) |  |
| Cold-wet | 111 (14.5) | 26 (8.6) | 85 (18.4) |  |
| Cold-dry | 67 (8.8) | 35 (11.6) | 32 (6.9) |  |
| **LVEF Phenotype, n (%)** |  |  |  | <0.001 |
| HFrEF | 469 (61.3) | 209 (69.2) | 260 (56.2) |  |
| HFmrEF | 104 (13.6) | 37 (12.3) | 67 (14.5) |  |
| HFpEF | 192 (25.1) | 56 (18.5) | 136 (29.4) |  |

n (%): Frequency (column percentage). IQR: interquartile range.
<sup>w</sup>: P-value from the Wilcoxon rank sum test of equality of medians. <sup>k</sup>: P-value from the Kruskal Wallis test of equality of medians. All other tests are from the Pearson's chi-square test.
LVEF: Left Ventricular Ejection Fraction. HFrEF: Heart Failure with reduced EF. HFmrEF: Heart Failure with mildly reduced EF. HFpEF: Heart Failure with preserved EF. NYHA: New York Heart Association. COPD: Chronic Obstructive Pulmonary Disease

**Figure 1:**
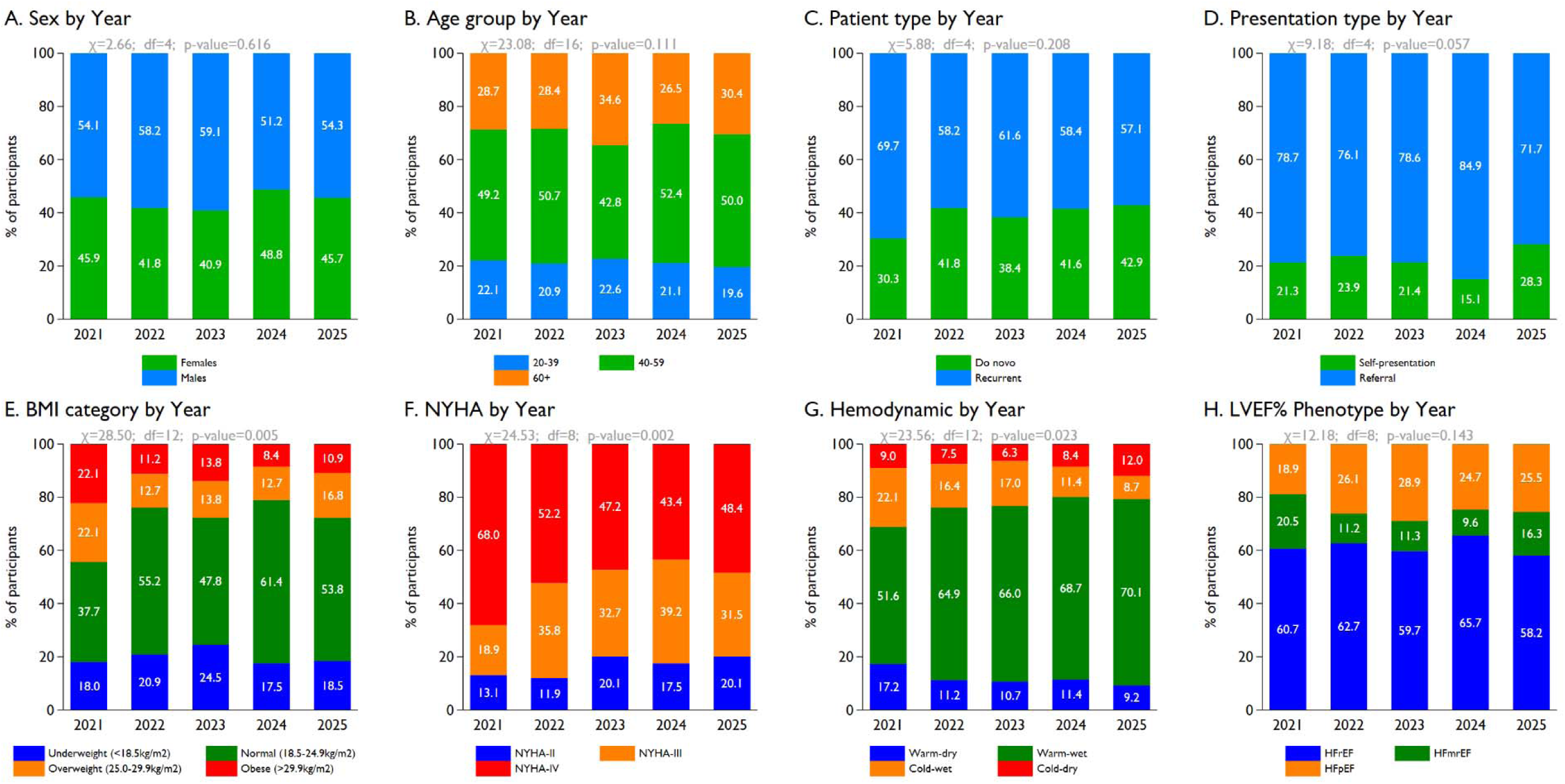
Trend of characteristics of heart failure patients by year of enrolment.

Systolic blood pressure (median 124 vs. 114 mmHg; p < 0.001), diastolic blood pressure (79 vs. 76 mmHg; p = 0.013), and mean arterial pressure (92 vs. 88 mmHg; p < 0.001) were significantly elevated in *de* novo patients, while heart rate was higher among recurrent cases (85 vs. 83 bpm; p = 0.005). No difference in BMI was observed between the two groups (p = 0.930), although approximately 19.9% of patients were underweight. While most patients (77.9%) were referred for further management, the referral rate was significantly higher for recurrent heart failure (89.6%) than for de novo heart failure (59.9%) (p < 0.001). (Table 1)

### 3.2 Aetiology of Heart Failure

The principal etiologies identified were dilated cardiomyopathy (DCM; 37.0%), hypertensive heart disease (HHD; 31.2%), valvular heart disease (VHD; 17.1%), ischemic heart disease (IHD; 6.3%), and other etiologies (7.1%). DCM, HHD, VHD, and IHD were each found to be significantly more prevalent among de novo patients, whereas “other aetiologies" were more common in recurrent cases (9.5% versus 3.3%; p = 0.001). (Table 1)

### 3.3. Cardiovascular Risk Factors and Comorbidities

Recurrent HF was strongly associated with a higher comorbidity burden. Risk factors like smoking (32.0% vs. 2.0%), family history of heart disease (31.1% vs. 11.3%), diabetes mellitus (32.4% vs. 4.0%), COPD (29.4% vs. 1.7%), and atrial fibrillation (20.7% vs. 11.3%) were all statistically significant among recurrent cases (all p < 0.001). Anaemia was also more common in recurrent HF (p = 0.033). The prevalence of hypertension (38.8% overall), CKD (25.8%), and stroke (9.5%) did not differ by patient type. Notably, 54.1% of patients had two or more comorbidities. (Table 1)

### 3.4. Clinical Severity at Presentation

On admission, a remarkable proportion of patients presented in an advanced functional class of NYHA IV (50.8%) and NYHA III (32.2%). Hemodynamic profiling revealed that two-thirds (65.1%) presented as warm-wet (congested but perfused), with cold-wet (cardiogenic shock-spectrum) profiles disproportionately seen in recurrent cases (18.4% vs. 8.6%; p < 0.001). HFrEF was the dominant phenotype (61.3%), followed by HFpEF (25.1%) and HFmrEF (13.6%). HFpEF was more prevalent in recurrent disease (29.4% vs. 18.5%; p < 0.001). (Table 1)

### 3.5. Sex- and Age-Stratified Patterns

Female patients had higher SBP, DBP, and MAP than males (all p ≤ 0.001) and a higher prevalence of HHD (35.1% vs. 28.1%; p = 0.039), VHD (20.8% vs. 14.2%; p = 0.016), DM (24.9% vs. 18.2%; p = 0.025), COPD (22.8% vs. 14.9%; p = 0.005), and atrial fibrillation (21.1% vs. 13.7%; p = 0.007). Anaemia was more common in males (30.3% vs. 21.3%; p = 0.005). (Table 2)

**Table 2:**
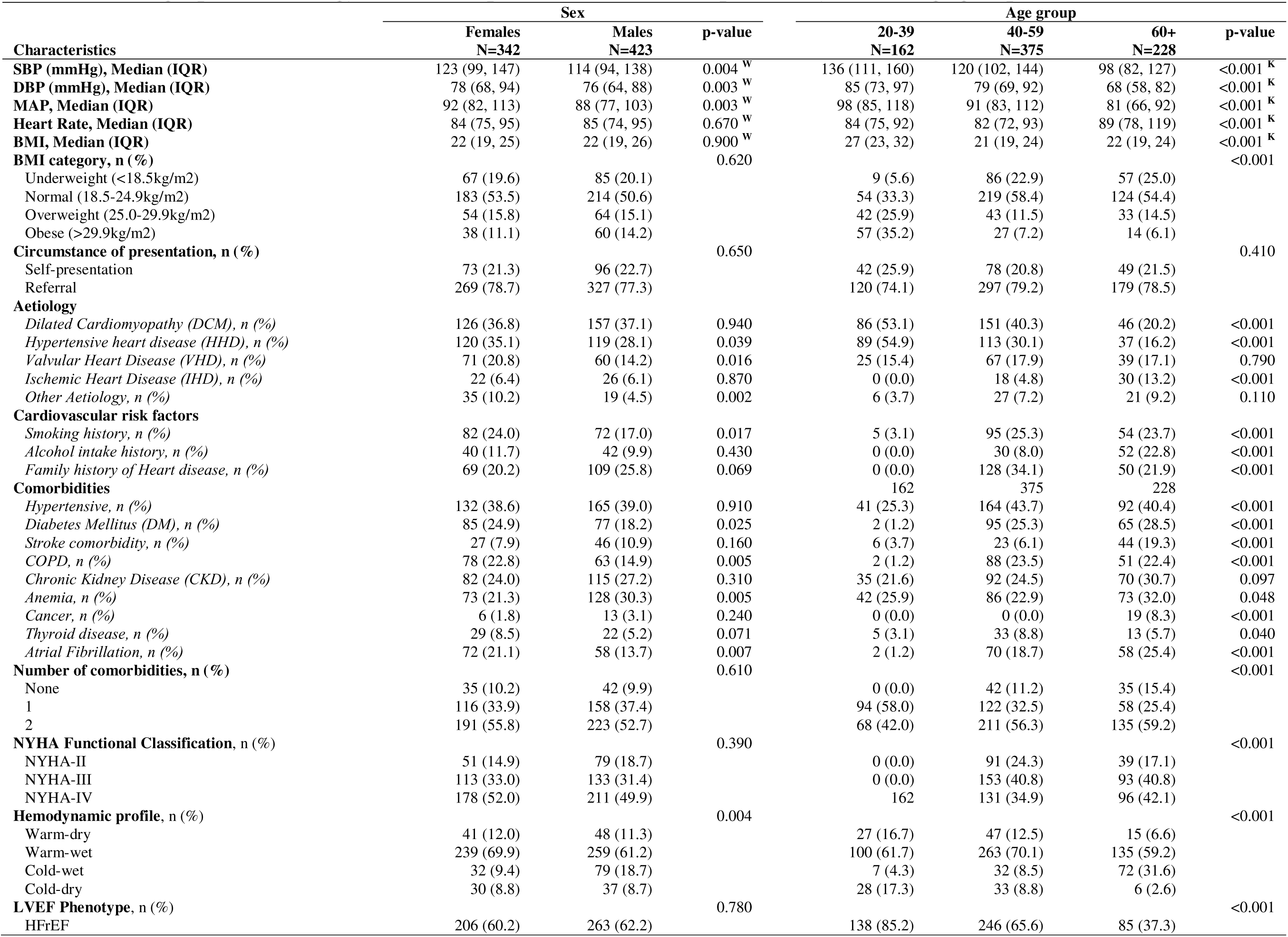
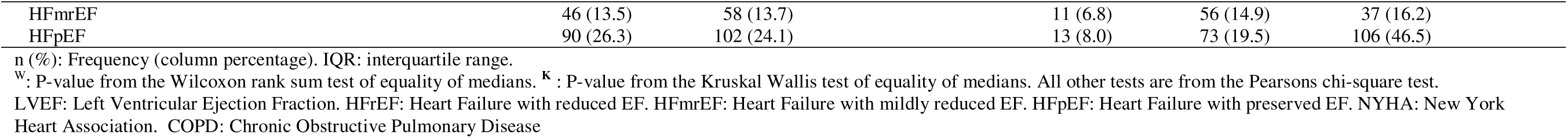
Demographics, Aetiology and clinical profile of heart failure patients by sex and age group.

Striking age variations were observed as blood pressure indices declined with age, while ischemic heart disease, stroke, atrial fibrillation, and HFpEF rose significantly. DCM and HHD were most prevalent in younger patients (20–39 years), accounting for 53.1% and 54.9% of cases, respectively. HFrEF dominated in younger adults (85.2%), whereas HFpEF predominated in those aged ≥60 years (46.5%). (Table 2)

### Temporal Trends, 2021–2025

Annual case volumes increased from 122 in 2021 to 184 in 2025, while demographic and aetiological distributions remained largely stable over the five-year period (all p-values> 0.05 for age, sex, aetiology, and most comorbidities). However, BMI category distribution shifted (p = 0.005), with declining proportions of overweight and obese patients over time. Clinically, NYHA class IV presentations declined from 68.0% in 2021 to 48.4% in 2025 (p = 0.002), and the cold-wet hemodynamic profile fell from 22.1% to 8.7% (p = 0.023). Prevalence of thyroid disease also declined significantly (p < 0.001). (Table 3)

**Table 3:** Demographics, Aetiology and clinical profile of heart failure patients by year of admission.

| Characteristics | Year of admission |  |  |  |  | P-value |
| --- | --- | --- | --- | --- | --- | --- |
|  | 2021<br>N=122 | 2022<br>N=134 | 2023<br>N=159 | 2024<br>N=166 | 2025<br>N=184 |  |
| **Age in years, Median (IQR)** | 52 (42, 61) | 55 (43, 62) | 53 (42, 61) | 53 (42, 60) | 54 (44, 61) | 0.800<sup>K</sup> |
| **Age group, n (%)** |  |  |  |  |  | 0.840 |
| 20-39 | 27 (22.1) | 28 (20.9) | 36 (22.6) | 35 (21.1) | 36 (19.6) |  |
| 40-59 | 60 (49.2) | 68 (50.7) | 68 (42.8) | 87 (52.4) | 92 (50.0) |  |
| 60+ | 35 (28.7) | 38 (28.4) | 55 (34.6) | 44 (26.5) | 56 (30.4) |  |
| **Sex, n (%)** |  |  |  |  |  | 0.620 |
| Females | 56 (45.9) | 56 (41.8) | 65 (40.9) | 81 (48.8) | 84 (45.7) |  |
| Males | 66 (54.1) | 78 (58.2) | 94 (59.1) | 85 (51.2) | 100 (54.3) |  |
| **SBP (mmHg), Median (IQR)** | 116 (92, 150) | 118 (95, 143) | 115 (97, 137) | 121 (100, 144) | 118 (99, 148) | 0.540<sup>K</sup> |
| **DBP (mmHg), Median (IQR)** | 79 (65, 95) | 76 (67, 92) | 76 (65, 87) | 78 (68, 92) | 77 (66, 92) | 0.560<sup>K</sup> |
| **MAP, Median (IQR)** | 91 (77, 116) | 89 (78, 110) | 88 (78, 100) | 92 (82, 112) | 89 (81, 114) | 0.550<sup>K</sup> |
| **Heart Rate, Median (IQR)** | 85 (75, 95) | 84 (78, 94) | 85 (73, 95) | 86 (76, 96) | 83 (73, 94) | 0.560<sup>K</sup> |
| **BMI, Median (IQR)** | 24 (20, 29) | 22 (19, 25) | 22 (19, 25) | 22 (20, 24) | 22 (19, 25) | 0.022<sup>K</sup> |
| **BMI category, n (%)** |  |  |  |  |  | 0.005 |
| Underweight (<18.5kg/m2) | 22 (18.0) | 28 (20.9) | 39 (24.5) | 29 (17.5) | 34 (18.5) |  |
| Normal (18.5-24.9kg/m2) | 46 (37.7) | 74 (55.2) | 76 (47.8) | 102 (61.4) | 99 (53.8) |  |
| Overweight (25.0-29.9kg/m2) | 27 (22.1) | 17 (12.7) | 22 (13.8) | 21 (12.7) | 31 (16.8) |  |
| Obese (>29.9kg/m2) | 27 (22.1) | 15 (11.2) | 22 (13.8) | 14 (8.4) | 20 (10.9) |  |
| **Circumstance of presentation, n (%)** |  |  |  |  |  | 0.057 |
| Self-presentation | 26 (21.3) | 32 (23.9) | 34 (21.4) | 25 (15.1) | 52 (28.3) |  |
| Referral | 96 (78.7) | 102 (76.1) | 125 (78.6) | 141 (84.9) | 132 (71.7) |  |
| **Aetiology** |  |  |  |  |  |  |
| Dilated Cardiomyopathy (DCM), n (%) | 49 (40.2) | 52 (38.8) | 54 (34.0) | 67 (40.4) | 61 (33.2) | 0.510 |
| Hypertensive heart disease (HHD), n (%) | 45 (36.9) | 42 (31.3) | 39 (24.5) | 57 (34.3) | 56 (30.4) | 0.200 |
| Valvular Heart Disease (VHD), n (%) | 16 (13.1) | 22 (16.4) | 34 (21.4) | 28 (16.9) | 31 (16.8) | 0.480 |
| Ischemic Heart Disease (IHD), n (%) | 4 (3.3) | 10 (7.5) | 10 (6.3) | 11 (6.6) | 13 (7.1) | 0.660 |
| Other Aetiology, n (%) | 10 (8.2) | 5 (3.7) | 12 (7.5) | 12 (7.2) | 15 (8.2) | 0.570 |
| **Cardiovascular risk factors** |  |  |  |  |  |  |
| Smoking history, n (%) | 29 (23.8) | 28 (20.9) | 29 (18.2) | 31 (18.7) | 37 (20.1) | 0.800 |
| Alcohol intake history, n (%) | 6 (4.9) | 17 (12.7) | 17 (10.7) | 19 (11.4) | 23 (12.5) | 0.240 |
| Family history of Heart disease, n (%) | 36 (29.5) | 33 (24.6) | 38 (23.9) | 33 (19.9) | 38 (20.7) | 0.330 |
| **Comorbidities** |  |  |  |  |  |  |
| Hypertensive, n (%) | 56 (45.9) | 46 (34.3) | 56 (35.2) | 62 (37.3) | 77 (41.8) | 0.240 |
| Diabetes Mellitus (DM), n (%) | 31 (25.4) | 29 (21.6) | 30 (18.9) | 32 (19.3) | 40 (21.7) | 0.690 |
| Stroke comorbidity, n (%) | 16 (13.1) | 14 (10.4) | 16 (10.1) | 16 (9.6) | 11 (6.0) | 0.320 |
| COPD, n (%) | 27 (22.1) | 26 (19.4) | 29 (18.2) | 27 (16.3) | 32 (17.4) | 0.760 |
| Chronic Kidney Disease (CKD), n (%) | 39 (32.0) | 31 (23.1) | 42 (26.4) | 40 (24.1) | 45 (24.5) | 0.500 |
| Anemia, n (%) | 39 (32.0) | 35 (26.1) | 47 (29.6) | 35 (21.1) | 45 (24.5) | 0.240 |
| Cancer, n (%) | 0 (0.0) | 6 (4.5) | 2 (1.3) | 3 (1.8) | 8 (4.3) | 0.055 |
| Thyroid disease, n (%) | 18 (14.8) | 10 (7.5) | 10 (6.3) | 8 (4.8) | 5 (2.7) | <0.001 |
| Atrial Fibrillation, n (%) | 18 (14.8) | 22 (16.4) | 22 (13.8) | 33 (19.9) | 35 (19.0) | 0.540 |
| **Number of comorbidities, n (%)** |  |  |  |  |  | 0.220 |
| None | 7 (5.7) | 14 (10.4) | 15 (9.4) | 24 (14.5) | 17 (9.2) |  |
| 1 | 38 (31.1) | 45 (33.6) | 57 (35.8) | 64 (38.6) | 70 (38.0) |  |
| 2 | 77 (63.1) | 75 (56.0) | 87 (54.7) | 78 (47.0) | 97 (52.7) |  |
| **NYHA Functional Classification, n (%)** |  |  |  |  |  | 0.002 |
| NYHA-II | 16 (13.1) | 16 (11.9) | 32 (20.1) | 29 (17.5) | 37 (20.1) |  |
| NYHA-III | 23 (18.9) | 48 (35.8) | 52 (32.7) | 65 (39.2) | 58 (31.5) |  |
| NYHA-IV | 83 (68.0) | 70 (52.2) | 75 (47.2) | 72 (43.4) | 89 (48.4) |  |
| **Hemodynamic profile, n (%)** |  |  |  |  |  | 0.023 |
| Warm-dry | 21 (17.2) | 15 (11.2) | 17 (10.7) | 19 (11.4) | 17 (9.2) |  |
| Warm-wet | 63 (51.6) | 87 (64.9) | 105 (66.0) | 114 (68.7) | 129 (70.1) |  |
| Cold-wet | 27 (22.1) | 22 (16.4) | 27 (17.0) | 19 (11.4) | 16 (8.7) |  |
| Cold-dry | 11 (9.0) | 10 (7.5) | 10 (6.3) | 14 (8.4) | 22 (12.0) |  |
| **LVEF Phenotype, n (%)** |  |  |  |  |  | 0.140 |
| HFrEF | 74 (60.7) | 84 (62.7) | 95 (59.7) | 109 (65.7) | 107 (58.2) |  |
| HFmrEF | 25 (20.5) | 15 (11.2) | 18 (11.3) | 16 (9.6) | 30 (16.3) |  |
| HFpEF | 23 (18.9) | 35 (26.1) | 46 (28.9) | 41 (24.7) | 47 (25.5) |  |

n (%): Frequency (column percentage). IQR: interquartile range.
<sup>w</sup>: P-value from the Wilcoxon rank sum test of equality of medians. <sup>k</sup>: P-value from the Kruskal Wallis test of equality of medians. All other tests are from the Pearson's chi-square test.
LVEF: Left Ventricular Ejection Fraction. HFrEF: Heart Failure with reduced EF. HFmrEF: Heart Failure with mildly reduced EF. HFpEF: Heart Failure with preserved EF. NYHA: New York Heart Association. COPD: Chronic Obstructive Pulmonary Disease

#### Factors associated with the aetiology of heart failure patients

Table 4 shows the bivariate associations between the five aetiologies of heart failure and patient characteristics.

**Table 4:**
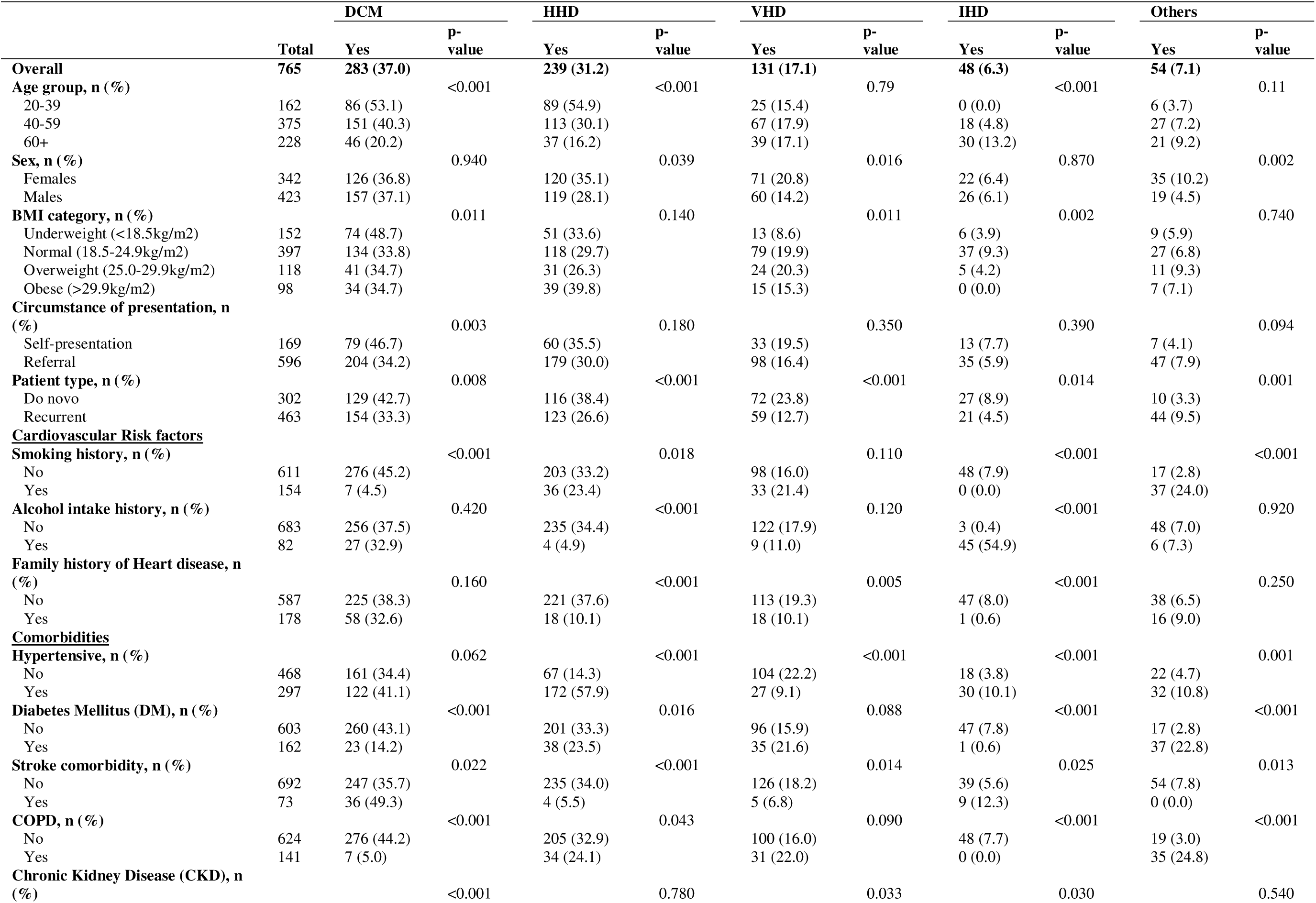
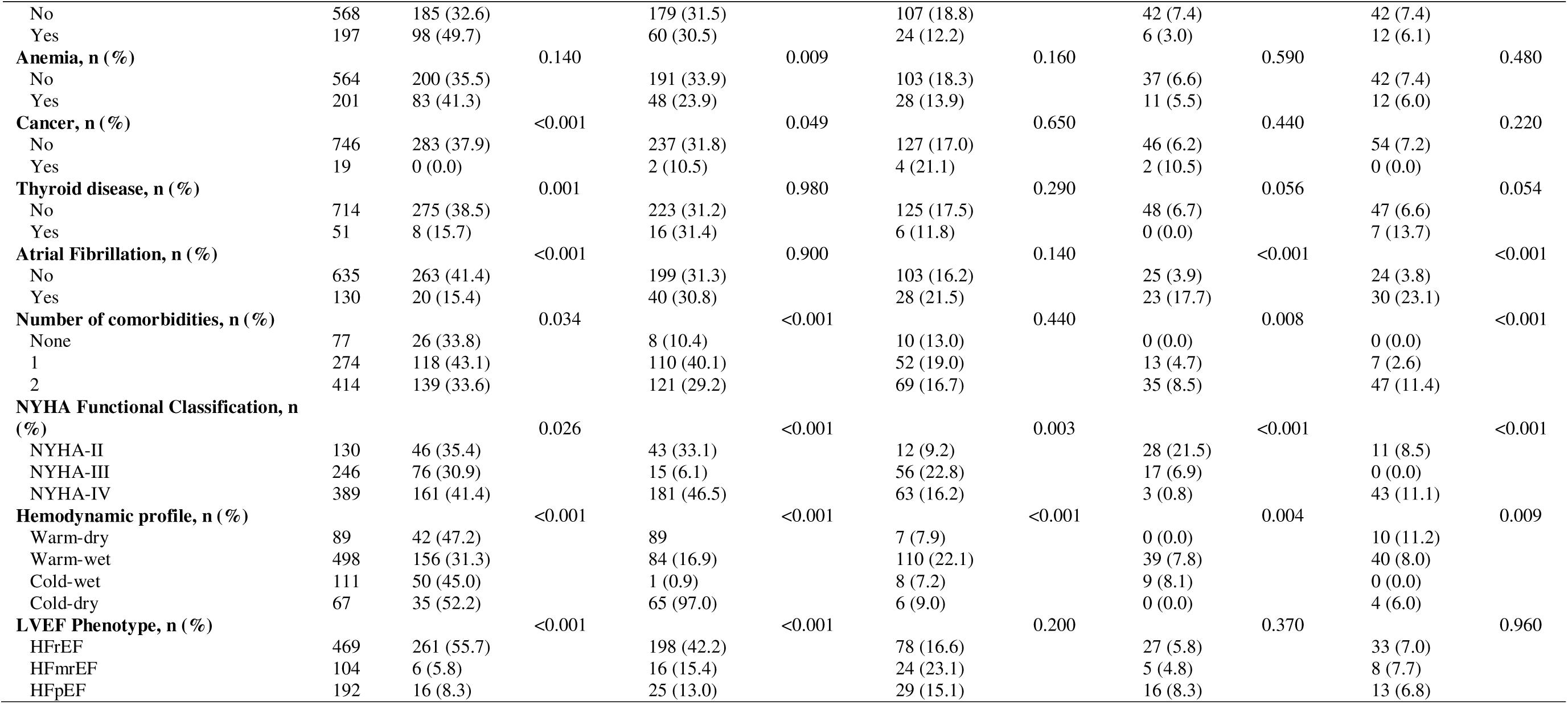
Bivariate association between characteristics and Aetiology of heart failure participants.

#### Multivariable Determinants of Aetiology

After adjustment for age, sex, BMI, presentation circumstance, patient type, year of admission, risk factors, and comorbidities, several independent associations emerged. DCM was inversely associated with older age (aOR: 0.13, 95% CI: 0.07–0.24 for ≥60 vs. 20–39 years; p < 0.001), higher BMI (aOR: 0.21, 95% CI: 0.11–0.43 for obese vs. underweight; p < 0.001), and COPD (aOR: 0.05, 95% CI: 0.02–0.16; p < 0.001), but positively associated with stroke (aOR: 2.05, 95% CI: 1.17–3.58; p = 0.012) and CKD (aOR: 2.21, 95% CI: 1.47–3.34; p < 0.001). HHD was associated with hypertension (aOR: 217.81, 95% CI: 73.13–648.76; p < 0.001) and atrial fibrillation (aOR: 11.63, 95% CI: 2.31–58.61; p = 0.003), while inversely associated with older age, higher BMI, recurrent presentation, alcohol use, and family history. (Table 5)

**Table 5:** Binary logistic regression model of factors associated with Aetiology among heart failure patients.

| Characteristics | Etiologies |  |  |  |  |  |  |  |  |  |
| --- | --- | --- | --- | --- | --- | --- | --- | --- | --- | --- |
|  | DCM |  | HHD |  | VHD |  | IHD |  | Others |  |
|  | aOR [95% CI] | P-value | aOR [95% CI] | P-value | aOR [95% CI] | P-value | aOR [95% CI] | P-value | aOR [95% CI] | P-value |
| **Age group** |  |  |  |  |  |  |  |  |  |  |
| 20-39 | 1.00 [reference] |  | 1.00 [reference] |  | 1.00 [reference] |  | 1.00 [reference]<sup>a</sup> |  | 1.00 [reference] |  |
| 40-59 | 0.57 [0.35, 0.93] | 0.024 | 0.07 [0.03, 0.15] | <0.001 | 2.16 [1.11, 4.20] | 0.024 | 1.00 [reference]<sup>a</sup> |  | 0.11 [0.02, 0.55] | 0.007 |
| 60+ | 0.13 [0.07, 0.24] | <0.001 | 0.05 [0.02, 0.14] | <0.001 | 2.27 [1.15, 4.51] | 0.019 | 2.34 [0.50, 11.08] | 0.283 | 0.09 [0.02, 0.46] | 0.004 |
| **Sex** |  |  |  |  |  |  |  |  |  |  |
| Females | 1.00 [reference] |  | 1.00 [reference] |  | 1.00 [reference] |  | 1.00 [reference] |  | 1.00 [reference] |  |
| Males | 0.95 [0.68, 1.34] | 0.775 | 0.64 [0.39, 1.05] | 0.080 | 0.60 [0.40, 0.90] | 0.014 | 1.66 [0.62, 4.43] | 0.313 | 0.55 [0.29, 1.04] | 0.067 |
| **BMI category** |  |  |  |  |  |  |  |  |  |  |
| Underweight (<18.5kg/m<sup>2</sup>) | 1.00 [reference] |  | 1.00 [reference] |  | 1.00 [reference] |  | 1.00 [reference] |  | 1.00 [reference] |  |
| Normal (18.5-24.9kg/m<sup>2</sup>) | 0.50 [0.31, 0.79] | 0.003 | 0.79 [0.41, 1.52] | 0.487 | 2.22 [1.18, 4.15] | 0.013 | 1.66 [0.40, 6.83] | 0.485 | 1.31 [0.48, 3.55] | 0.595 |
| Overweight (25.0-29.9kg/m<sup>2</sup>) | 0.34 [0.18, 0.64] | 0.001 | 0.31 [0.13, 0.74] | 0.008 | 2.52 [1.16, 5.46] | 0.019 | 2.00 [0.27, 14.79]<sup>b</sup> | 0.499 | 2.55 [0.77, 8.41] | 0.124 |
| Obese (>29.9kg/m<sup>2</sup>) | 0.21 [0.11, 0.43] | <0.001 | 0.29 [0.11, 0.73] | 0.009 | 2.39 [1.02, 5.60] | 0.046 | 2.00 [0.27, 14.79]<sup>b</sup> | 0.499 | 3.29 [0.90, 12.00] | 0.071 |
| **Circumstance of presentation** |  |  |  |  |  |  |  |  |  |  |
| Self-presentation | 1.00 [reference] |  | 1.00 [reference] |  | 1.00 [reference] |  | 1.00 [reference] |  | 1.00 [reference] |  |
| Referral | 0.58 [0.38, 0.87] | 0.009 | 1.14 [0.64, 2.00] | 0.660 | 0.93 [0.53, 1.65] | 0.812 | 0.63 [0.16, 2.45] | 0.504 | 0.64 [0.17, 2.34] | 0.497 |
| **Patient type** |  |  |  |  |  |  |  |  |  |  |
| Do novo | 1.00 [reference] |  | 1.00 [reference] |  | 1.00 [reference] |  | 1.00 [reference] |  | 1.00 [reference] |  |
| Recurrent | 1.39 [0.95, 2.03] | 0.088 | 0.11 [0.05, 0.23] | <0.001 | 0.36 [0.21, 0.60] | <0.001 | 3.74 [1.11, 12.54] | 0.033 | 1.19 [0.38, 3.78] | 0.766 |
| **Year of admission** |  |  |  |  |  |  |  |  |  |  |
| 2021 | 1.00 [reference] |  | 1.00 [reference] |  | 1.00 [reference] |  | 1.00 [reference] |  | 1.00 [reference] |  |
| 2022 | 0.86 [0.47, 1.58] | 0.628 | 1.12 [0.44, 2.87] | 0.817 | 0.98 [0.47, 2.01] | 0.947 | 1.08 [0.17, 6.78] | 0.936 | 0.40 [0.12, 1.33] | 0.136 |
| 2023 | 0.69 [0.39, 1.22] | 0.197 | 0.68 [0.29, 1.62] | 0.385 | 1.57 [0.78, 3.13] | 0.204 | 0.76 [0.14, 4.07] | 0.744 | 1.12 [0.38, 3.30] | 0.838 |
| 2024 | 0.89 [0.50, 1.59] | 0.689 | 1.11 [0.43, 2.88] | 0.823 | 0.96 [0.47, 1.96] | 0.904 | 1.15 [0.18, 7.32] | 0.884 | 0.91 [0.39, 2.16] | 0.838 |
| 2025 | 0.62 [0.35, 1.09] | 0.098 | 0.52 [0.21, 1.28] | 0.154 | 0.93 [0.45, 1.91] | 0.844 | 0.82 [0.17, 4.02] | 0.802 | 1.20 [0.48, 3.01] | 0.691 |
| **Alcohol intake** | 1.14 [0.57, 2.26] | 0.709 | 0.00 [0.00, 0.00] | <0.001 | 0.49 [0.20, 1.20] | 0.117 | - | - | 7.02 [1.33, 36.99] | 0.022 |
| **Family history of heart disease** | 0.76 [0.48, 1.20] | 0.235 | 0.06 [0.02, 0.17] | <0.001 | 0.55 [0.26, 1.14] | 0.107 | 0.20 [0.02, 1.71] | 0.141 | 10.17 [2.47, 41.92] | 0.001 |
| **comorbidities** |  |  |  |  |  |  |  |  |  |  |
| Hypertension | 1.01 [0.70, 1.45] | 0.970 | 17.81 [3.13, 48.76] | <0.001 | 0.36 [0.22, 0.61] | <0.001 | 6.80 [2.00, 23.18] | 0.002 | 5.74 [2.62, 12.56] | <0.001 |
| Diabetes Mellitus | 1.52 [0.62, 3.71] | 0.357 | 0.07 [0.01, 0.77] | 0.030 | 3.01 [0.96, 9.48] | 0.059 | 0.07 [0.00, 9.10] | 0.289 | 6.33 [1.30, 30.91] | 0.022 |
| Stroke | 2.05 [1.17, 3.58] | 0.012 | 0.07 [0.01, 0.37] | 0.002 | 0.41 [0.14, 1.19] | 0.100 | - | - | - | - |
| COPD | 0.05 [0.02, 0.16] | <0.001 | 2.16 [0.23, 20.49] | 0.503 | 0.71 [0.23, 2.24] | 0.560 | 3.22 [0.37, 28.41] | 0.292 | 5.24 [1.05, 26.05] | 0.043 |
| Chronic kidney disease | 2.21 [1.47, 3.34] | <0.001 | 0.55 [0.32, 0.94] | 0.029 | 0.79 [0.47, 1.33] | 0.375 | 0.41 [0.10, 1.68] | 0.214 | 0.95 [0.46, 1.94] | 0.888 |
| Anemia | 1.30 [0.87, 1.95] | 0.203 | 0.55 [0.32, 0.93] | 0.026 | 0.83 [0.50, 1.38] | 0.470 | 3.08 [1.00, 9.53] | 0.051 | 0.77 [0.35, 1.71] | 0.526 |
| Cancer | - | - | 0.51 [0.16, 1.61] | 0.253 | 0.99 [0.29, 3.37] | 0.993 | 0.30 [0.03, 2.80] | 0.288 | - | - |
| Thyroid disease | 0.54 [0.20, 1.42] | 0.212 | 3.20 [1.13, 9.05] | 0.028 | 0.41 [0.17, 1.02] | 0.055 | - | - | 0.79 [0.32, 1.97] | 0.615 |
| Atrial Fibrillation | 0.93 [0.44, 1.99] | 0.855 | 11.63 [2.31, 58.61] | 0.003 | 1.12 [0.48, 2.63] | 0.795 | 1.75 [0.35, 8.79] | 0.494 | 4.91 [0.95, 25.45] | 0.058 |

aOR: adjusted odds ratio. CI: confidence interval DCM: Dilated Cardiomyopathy. HHD: Hypertensive heart disease. VHD: Valvular Heart Disease. IHD: Ischemic Heart Disease
<sup>a</sup>: age group 20-39 and 40-59 were combined as reference. <sup>b</sup>: overweight and obese were combined as one category

VHD was more likely in older patients (aOR: 2.27, 95% CI: 1.15–4.51 for ≥60 years), in those with normal-to-obese BMI, and was less common in males (aOR: 0.60; p = 0.014), recurrent patients (aOR: 0.36; p < 0.001), and those with hypertension. IHD was independently associated with hypertension (aOR: 6.80, 95% CI: 2.00–23.18; p = 0.002) and recurrent HF (aOR: 3.74, 95% CI: 1.11–12.54; p = 0.033). Other aetiologies were strongly linked to alcohol use (aOR: 7.02), family history (aOR: 10.17), hypertension (aOR: 5.74), DM (aOR: 6.33), and COPD (aOR: 5.24). (Table 5)

## 4.0 Discussion

This report presents the first in-depth analysis of hospitalized patients with heart failure in Sierra Leone. Our data indicate that the acute presentation of heart failure constitutes 10% of all medical admissions to the Intensive Care Unit in the hospital. Cardiovascular conditions constitute approximately twenty percent of all medical emergency admissions, ranking as the second most common cause following infections and infestations, which comprise nearly fifty percent of all cases.

In our study, young and middle-aged persons were primarily impacted by heart failure, with an average age of 53 years. About half (49%) of the affected population in the age group of 40–59 years. This illustrates the early onset of cardiovascular disease in our population, resulting in a considerable loss of productive years and significant socioeconomic consequences. Our findings are consistent with reports from other Sub-Saharan African HF registries, such as THESUS-HF [7] and INTER-CHF [2], which also documented average ages in the early 50s.

From the OPTIMIZE-HF and ADHERE registries in the United States, older populations with median ages of 65 and 70 years, respectively, were reported, which is contrary to our study. Furthermore, the ESC Heart Failure Long-Term Registry in Europe reports a mean age of 67 years [17]. This difference accentuates the unique epidemiology of Sub-Saharan Africa, characterized by early-onset heart failure mainly due to hypertensive heart disease, rheumatic heart disease (RHD), and HIV-associated cardiomyopathy [7]. Limited preventive approaches, delayed diagnoses, and healthcare barriers contribute additionally to early disease presentation in Sub-Saharan Africa.

At the time of presentation, more females reported having a significantly higher incidence of hypertension, heart failure with preserved ejection fraction (HFpEF), atrial remodelling, diabetes, and valvular disease, while males demonstrated more severe manifestations of cardiac dysfunction, which was accompanied by anaemia and greater ventricular impairment. These sex-related disparities have been previously documented and are likely attributable to a multifactorial interplay involving cardiovascular risk factors, sociocultural influences, hormonal effects, medical care access, and variations in heart failure phenotypes between genders in Sub-Saharan Africa [18-21].

The prevalence of acute heart failure phenotypes was 61.3% for HFrEF, 13.6% for HFmrEF, and 25.1% for HFpEF in our study. This is consistent with previous studies, which demonstrated HFrEF prevalence ranging from 36% to 66%, HFmrEF from 14% to 30%, and HFpEF from 16% to 43% [22-24]. The INTER-CHF study [2] reported 53.7% HFpEF, 30.1% HFmrEF, and 16.2% HFrEF, while the Swedish Heart Failure Registry reported 56% HFrEF, 21% HFmrEF, and 23% HFpEF [25]. A multicentre study in Japan indicated 36% HFrEF, 21% HFmrEF, and 43% HFpEF [26]. Differences between these studies and ours may arise from variations in methodology, emphasis on acute heart failure, shifts in cardiovascular risk profiles, and the younger age of our population [27-29].

The most common underlying aetiologies of heart failure in the study cohort were cardiomyopathies (37.0%) and hypertensive heart diseases (31.2%), while other aetiologies were rheumatic valvular (17.1%) and ischemic heart disease (6.3%). These findings, consistent with previous reports from several West African nations, may reflect the emerging adoption of Western dietary and lifestyle habits across the subregion [9,10,12, 30-32]. In sub-Saharan Africa, multiple key causes of dilated cardiomyopathy have been identified, including HIV-related cardiomyopathy, peripartum cardiomyopathy, myocarditis, alcohol-induced cardiomyopathy, and genetic forms [33-35]. Familial disease accounts for 30–50% of cases, depending on the extent of the investigation [34]. The current study found idiopathic dilated cardiomyopathy to be the most common form, at 85.3%, compared with 65.1% reported by Agyekum et al and 50% reported by Felker et al [9,35]. The high rate of idiopathic cases observed in our study may be influenced by methodological limitations, including incomplete investigations, as echocardiography is not very sensitive for distinguishing causes related to dilated cardiomyopathy.

The absence of cardiac MRI and coronary angiography, due to resource scarcities in Sierra Leone, further restricted our capability to precisely determine underlying aetiologies. Consequently, the reported proportions should be interpreted with caution as they may reflect these diagnostic limitations rather than the true epidemiological distribution.

Hypertensive heart disease is the most common form of heart failure as reported by various African heart failure registries, including THESUS-HF [7] and INTER-CHF [2]. This pattern is consistent with our findings yet contrasts with those observed in high-income regions such as North America, Latin America and Europe, where ischemic heart disease remains the primary aetiology [36-38]. In Sierra Leone, the detection, awareness, and management of hypertension are considerably hindered by restricted accessibility of medical services, medication non-adherence, poverty, and illiteracy [39]. Our data revealed significantly higher blood pressure and mean arterial pressure (MAP) in the “de novo HF patients” compared to “recurrent HF patients."

Blood pressure was either within the normal range or reduced in some cases of recurrent heart failure, a phenomenon termed as “burnt-out hypertension”. This is associated with advanced cardiac disease, showing a poor cardiac reserve rather than the absence of hypertension [10,40,41]. Consequently, the absence of elevated blood pressure at presentation does not necessarily rule out hypertension as the underlying aetiology of heart failure, especially within the African context.

Consistent with reports from neighbouring countries, degenerative valvular heart disease, rather than rheumatic valve disease, was reported as the leading aetiology of valvular heart failure, and this is similar to the 17.9% of valvular heart failure cases reported in our study [9,32,41]. A greater burden of comorbidities and risk factors among patients with recurrent heart failure in comparison to de novo patients was reported in our study. These included smoking (32.0% vs. 2.0%), diabetes (32.4% vs. 4.0%), chronic obstructive pulmonary disease (COPD) (29.4% vs. 1.7%), atrial fibrillation (20.7% vs. 11.3%), and family history (31.1% vs. 11.3%) (all p < 0.001). Reports from Africa have documented similar events and are related to inadequate diagnostic procedures and insufficient availability of comprehensive long-term cardiovascular care. This illustrates the complex multisystemic nature of heart failure [4,7,10,21,24,30].

Most of the patients in our study diagnosed with advanced heart failure exhibited a warm-wet hemodynamic profile, indicative of extensive pulmonary congestion at presentation. Furthermore, most patients with advanced heart failure demonstrated recurrent episodes, which may suggest late referral and poor management of heart failure in Sierra Leone. Our findings were similar to those of the THESUS-HF registry [7] and other African studies, in which late-stage HF presentation was prevalent. [9,41,42,43]. In Western nations, the diagnosis of heart failure is made at an earlier stage of the disease. This is due to routine screening, medical care accessibility and cardiovascular risk factors management. Establishment of HF screening programs and the decentralisation of HF clinics in SSA are probable strategies to deal with these gaps and increase clinical outcomes.

Over the five-year study period, a pronounced trend in heart failure (HF) admissions was identified, characterised by a consistent increase in the number of cases admitted. This pattern may indicate either a true increase in burden or an enhancement in diagnostic awareness. The trend could also be attributable to the rising prevalence of cardiovascular risk factors, such as hypertension, diabetes, and obesity, within Sierra Leone [39,44,45]. The increased detection of heart failure cases is likely due to clinician awareness, refined diagnostic procedures and hospital referral.

Despite the increase in admissions, several clinical parameters including body mass index (BMI), advanced heart failure symptoms (NYHA functional class IV), and the cold-wet hemodynamic profile have declined. This reduction may reflect earlier recognition of heart failure severity, improved health-seeking behaviour, and more timely clinical interventions. Furthermore, the decrease in the cold-wet hemodynamic profile suggests progress in heart failure management strategies.

After adjusting for demographic and clinical variables, several independent predictors of heart failure phenotypes were further identified. Dilated cardiomyopathy (DCM) was found to be more prevalent among younger patients and exhibited a positive correlation with higher body mass index (BMI), chronic obstructive pulmonary disease (COPD), stroke, and chronic kidney disease. This indicates the important role of obesity, multisystem comorbidities, and chronic inflammatory states in myocardial dysfunction [46,47]. These data are consistent with past research conducted in Africa, which illustrates that non-ischemic cardiomyopathies predominantly affect younger populations in sub-Saharan Africa [48,49].

There was a strong correlation of hypertensive heart disease with increasing age and the presence of atrial fibrillation. This association illustrates the long-term consequences of poorly managed hypertension, which contributes to both structural and electrical remodelling of the heart [41,46].

The presence of atrial fibrillation further worsens cardiac dysfunction and heightens the risk of thromboembolic events. In general, these data demonstrate the complex nature of HF in Africa, stressing the significance of early intervention in cardiovascular risks, improved management of hypertension and the adoption of integrated multidisciplinary care strategies. A significant nutritional paradox of frailty is apparent in our study, with 19.9% of patients being underweight, illustrating the combined burden of undernutrition and cardiovascular disease in this resource-constrained environment. A similar finding was documented by Manza et al, wherein there is an association between frailty and the outcomes of heart failure hospitalization [50].

### 4.1 Clinical Implications of the findings

Our study has provided significant information for clinical practice and public health implementation in SSA. Based on the high prevalence of heart failure among young adults and the common etiologies identified, clinicians as well as policymakers should prioritize the development and implementation of targeted screening procedures for high-risk populations in Sierra Leone. Early detection programs at primary care levels, routine cardiovascular risk assessment for persons under 60 years and strengthening referral pathways for timely specialist care are recommended. These measures are likely to facilitate earlier diagnosis, improve clinical outcomes, and strengthen healthcare efficiency throughout various medical facilities in Sierra Leone.

### 4.2 Strengths of the study

This study represents the first single-centre investigation of heart failure (HF) in Sierra Leone, granting essential epidemiological and clinical findings of a younger patient population, a predominance of hypertensive HF, dilated cardiomyopathy, and frequent presentation with advanced symptoms. The findings from this study provide a framework for strengthening primary health care by promoting early HF detections, improving access to affordable diagnostics, integrating HF management into chronic disease programs and adopting digital health solutions such as telemedicine.

### 4.3 Limitations of the study

The limitations identified in the study prevent its generalizability, as it is a hospital-based, retrospective design conducted at a single tertiary centre. The findings may primarily reflect the characteristics and healthcare-seeking patterns of patients accessing advanced facilities in an urban setting rather than those of the rural communities, who may experience barriers to tertiary care. As an observational study, the risks of selection bias, underreporting or miscoding of comorbidities and outcomes are inherent. These variables collectively limit the extent to which the study’s conclusions can be extrapolated beyond the studied population.

### 4.4 Conclusion

This study provided useful information on the epidemiology, clinical characteristics and temporal trends of heart failure (HF) in Sierra Leone. The findings show a younger patient population, a predominance of hypertensive heart failure, dilated cardiomyopathy and advanced symptoms severity at presentation. This pattern is different from industrialized nations and may suggest systemic deficiencies in Sierra Leone’s health service delivery. These challenges will therefore necessitate an investment in encouraging public awareness of cardiovascular diseases, upgrading diagnostic capacity and increasing access to evidence-based therapies. Furthermore, establishing long-term registries of heart failure and other cardiovascular diseases is imperative for monitoring trends and informing health policy.

## Acknowledgements

We sincerely acknowledge the support of Choithrams Memorial Hospital in granting the researchers access to its digital medical platform to conduct this retrospective study.

## Contributors

JBWR conceived the study idea, drafted and revised the manuscript, and is the guarantor. MS contributed to data collection and revised the manuscript. YA contributed to the data analysis and revised the manuscript. JMC, ET, NB, KB, MK, OZM, and DRL reviewed the manuscript. All co-authors reviewed and approved the final manuscript.

## Patient consent for publication

Not applicable.

## Ethics approval

The study was approved by the Sierra Leone Ethical and Review Committee.

## Provenance and peer review

Not commissioned; internally peer-reviewed.

## Data availability statement

Data are available upon reasonable request. Anonymised data are available upon reasonable request to the corresponding author.

